# Online46: online cognitive assessments in elderly cohorts - the British 1946 birth cohort case study

**DOI:** 10.1101/2024.09.19.24313984

**Authors:** Ziyuan Cai, Valentina Giunchiglia, Rebecca Street, Martina Del Giovane, Kirsty Lu, Maria Popham, Andrew Wong, Heidi Murray-Smith, Marcus Richards, Sebastian Crutch, Peter J. Hellyer, Jonathan M Schott, Adam Hampshire

## Abstract

**INTRODUCTION:** Online assessments are scalable and cost-effective for detecting cognitive changes, especially in elderly cohorts with limited mobility and higher vulnerability to neurological conditions. However, determining the uptake, adherence, and usability of these assessments in older adults, who may have less experience with mobile devices is crucial.

**METHODS:** 1,776 members (aged 77) of the MRC National Survey of Health and Development (NSHD) were invited to complete 13 online cognitive tasks. Adherence was measured through task compliance, while uptake (consent, attempt, completion) was linked to health and sociodemographic factors. Usability was evaluated through qualitative feedback.

**RESULTS:** This study’s consent (56.9%), attempt (80.5%), and completion (88.8%) rates are comparable to supervised NSHD sub-studies. Significant predictors of uptake included education, sex, handedness, cognitive scores, weight, smoking, alcohol consumption, and disease burden.

**DISCUSSION:** With key recommendations followed, online cognitive assessments are feasible, with good adherence, and usability in older adults.

## 1. BACKGROUND

Cognitive tasks are crucial for detecting and monitoring age-related decline. Traditionally, these assessments are conducted in pen-and-paper format under supervised conditions, tailored to specific cognitive domains and clinical populations [1]. Automated online assessments offer an alternative approach with several advantages. They can match or surpass popular pen-and-paper tasks in detecting cognitive impairments [2–5], reduce the burden on patients and clinical staff since patients can conduct assessments in any unsupervised environment [6], and enable cost- and time-effective large-scale longitudinal data collection.

Online assessments are particularly beneficial for older adults, who may struggle to attend clinics in-person. However, technology anxiety and limited access to devices may pose challenges to engagement [7–9], making it essential to determine the uptake of online testing technology in this age range. Additionally, unsupervised cognitive tasks may introduce data errors due to the lack of support, necessitating robust investigation of adherence, usability and sampling bias.

Previous studies have focused on assessing the validity of online assessments by measuring their ability to discriminate clinical conditions [10–12] and by benchmarking their performance compared to standard pen-and-paper tasks [13] and computerised tasks conducted in supervised settings [14]. However, they typically lack detailed analysis of factors influencing adherence to cognitive assessments or are not focused on general ageing populations [15]. Here, we examine the application of computerised cognitive assessments in an elderly cohort (age=77) from the MRC National Survey of Health and Development. Our first aim was to assess uptake when recruiting an older adult cohort to an online assessment and to determine the factors that predict the probability of participation and completion. Our second aim was to assess patterns of adherence and compliance. Finally, we aimed to qualitatively analyse information on usability and barriers for participation, to provide recommendations for future protocol optimisation.

## 2. METHODS

### 2.1 The 1946 British Birth Cohort Study Sample

The MRC National Survey of Health and Development (NSHD) is the longest continually studied British birth cohort, comprising 5362 individuals born in England, Scotland, and Wales during the first week of March 1946 [16]. The study has collected sociodemographic, medical, cognitive and psychological function data through interviews and examinations in 27 waves, as well as smaller sub-study collections, approximately 2700 participants in active follow-up [17]. The current aim of the NSHD study is to explore long-term ageing and how it is affected by factors across the life course [18–21].

### 2.2 Study Recruitment

This study, called Online46, assessed the uptake, adherence and usability of automated online assessments in the NSHD cohort in 2023. It involved a pilot stage to design the assessments and a main stage for large-scale data collection. Twenty-three study members from the participant advisory panel piloted the study. The main stage had a soft launch with a small actively engaged subset of participants (N=50), and then a full launch a week later with the remaining (N=1703) participants (Figure 1). Participants had 4 weeks to complete the online assessments. Follow-up email invitations were sent after 2 and 3.5 weeks.

**Figure 1.**
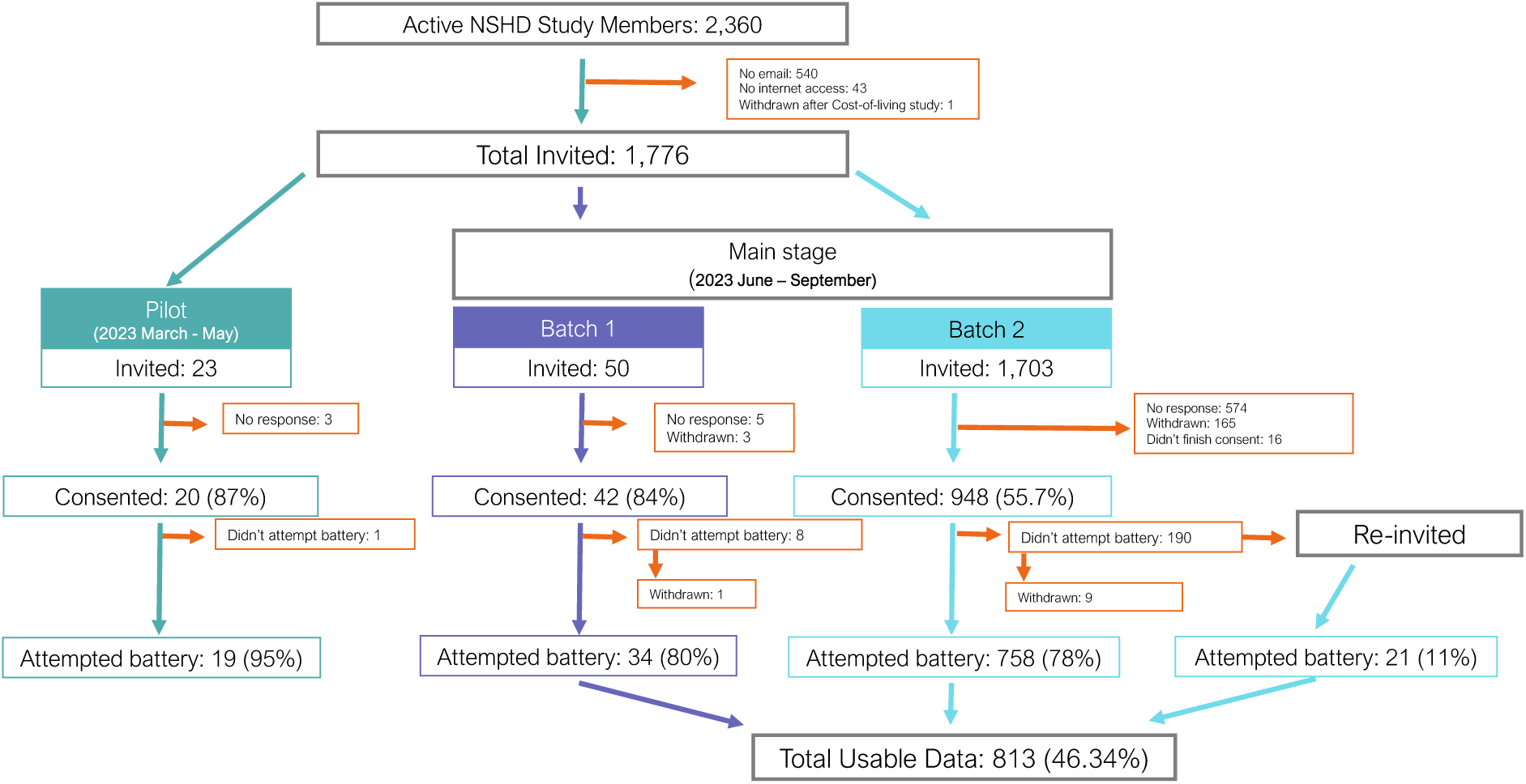
Recruitment flowchart for the Online46 study. Out of 2,360 active NSHD members, 1776 were invited to participate in the Online46 study. 1010 members consented and 813 of them attempted the online cognitive battery. The study involved two stages: 1) a pilot stage between March and May for assessment design and 2) a main stage between June and September for data collection.

We recruited from among all active NSHD study members (N=2360) excluding those with no email (N=540), no internet access (N=43) or who had withdrawn during previous data collection (N=1). 1776 members were invited through emails. Participants completed consent forms via a Qualtrics link and received a user-specific link to the cognitive assessment. After 4 weeks, non-attempting participants (N=190) were re-invited and were given an extra week to attempt the battery.

### 2.3 Cognitive task battery

Cognitron is a server system used for online cognitive assessments. It hosts more than 100 optimised cognitive tasks that are sensitive, domain specific and widely validated in general population and clinical cohorts [2, 11, 22–25]. More information is available on the website https://www.cognitron.co.uk/.

Here, participants were asked to undertake 13 Cognitron tasks selected to measure different aspects of reaction time, motor control, memory, attention, reasoning and executive functions (Figure 2, Supplementary Table 1). Participants could access the tasks through web browsers on any smartphone, tablet or personal computer. The tasks were presented as a battery in fixed order without supervision. General instructions were provided at the beginning of the battery. Specific instructions were presented before each task followed by a brief set of practice trials. Information related to performance (accuracy and reaction time), compliance (e.g., time spent outside of the task webpage) and detailed trial-by-trial responses were collected automatically.

**Figure 2.**
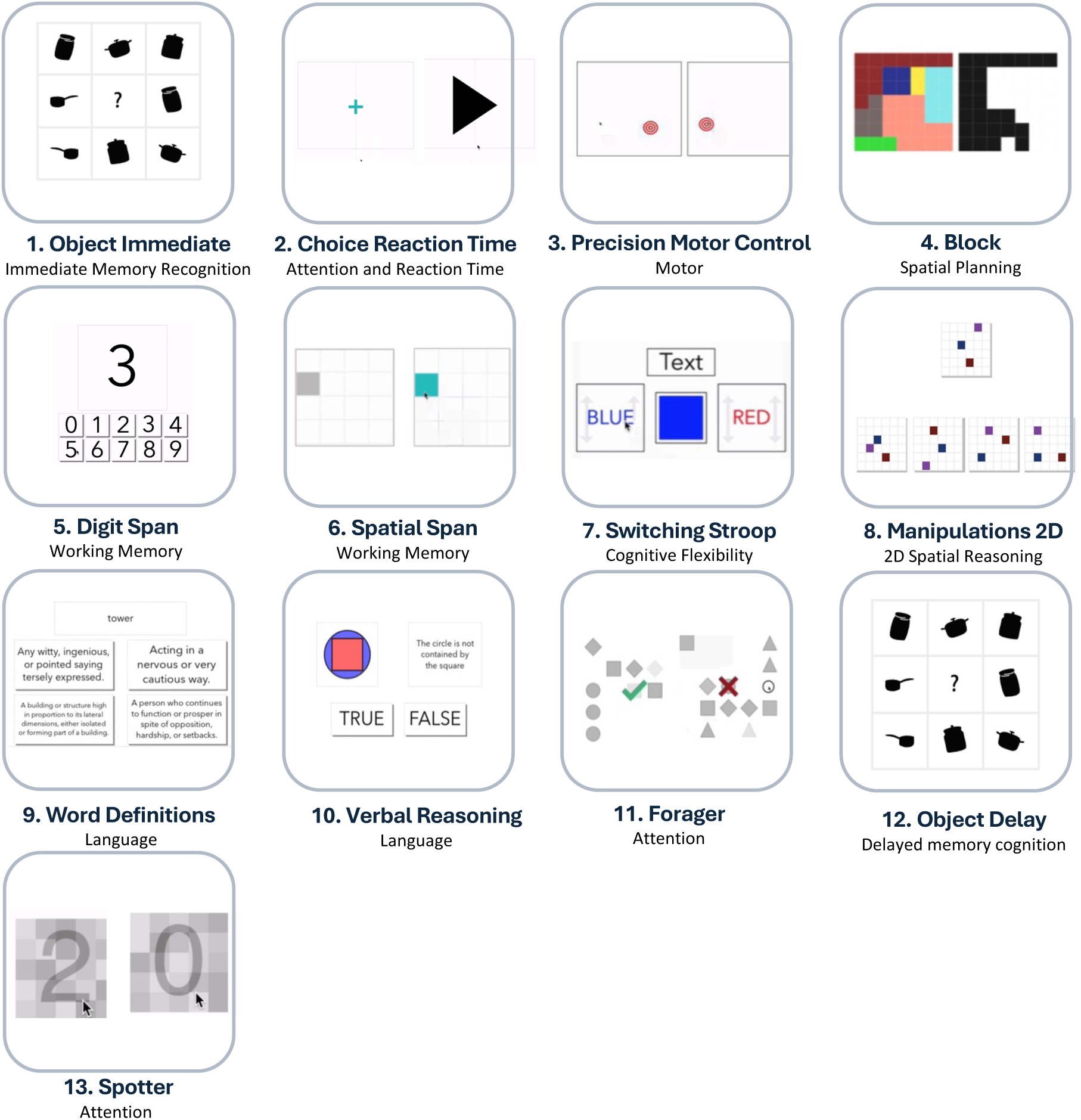
Schematic of the cognitive assessment. Participants were presented with 13 cognitive tasks (listed in order with corresponding cognitive domain): Object Memory Immediate (immediate recognition memory), Choice Reaction Time (attention and reaction time), Precision Motor Control (motor), Blocks (Spatial Planning), Digit Span (working memory capacity), Spatial Span (working memory capacity), Switching Stroop (attentional control), Manipulations 2D (2D spatial reasoning), Word Definition (crystalised knowledge of words), Verbal Reasoning (grammar-based reasoning), Forager (contingency reversal learning), Object delay (delayed recognition memory) and Spotter (sustained attention).

### 2.4 Uptake

We investigated how sociodemographic and health-related characteristics differed at three levels of recruitment: 1) consented to take part, 2) attempted any of the tasks, and 3) completion of the entire online assessment. The sociodemographic and health-related characteristics of the NSHD cohort have been described in previous publications [20] and are summarised in Supplementary Table 2.

Three multivariable logistic regression models were conducted to predict consent and battery attempt and completion, using the same set of independent variables: socio-demographic factors, mental health, smoking history, alcohol consumption, BMI, disease burden and self-rated health. Statistical significance was assessed using a Wald test after fitting the models. Multivariable regression was used to align with previous NSHD recruitment studies [20], and control for confounding variables.

Whether the accuracy score on the first task (Object Memory Immediate) predicted the likelihood of completing the entire assessment was evaluated with a logistic regression model.

### 2.5 Adherence

#### 2.5.1 Task compliance

Based on the task design, different measures of compliance were assessed. Conservative criteria were applied to prevent from removing clinically relevant data. For Choice Reaction Time (CRT) and Word Definitions, the number of time-out trials, defined as trials in which participants do not interact with the stimulus, was calculated. Participants with at least 90% of trials being defined as time-out trials were marked as not complying. For Verbal Reasoning, Manipulation 2D, Switching Stroop, CRT and Object Immediate and Delay, instances of clicking repetitively in the same screen location were measured. Participants who pressed in the same location in all trials of a given task were defined as not engaged. For all tasks, participants who missed the stimulus in all trials were considered unengaged. Finally, participants with a summary score below a set expected threshold were also defined as non-compliant as this can be a sign that the task was not completed properly. These thresholds were determined for each task using an automated method that processes the histogram distributions and compares the bin heights against predefined thresholds (i.e., the 5th percentile of the left tail of the distribution). Details in Supplementary Text 1.

#### 2.5.2 Cheating

Digit Span requires participants to memorise a sequence of digits, whose length increases by one digit at each trial. A small proportion of participants write the digits in another browser window to move forward in the assessment. Participants within the top 10% quantile of the Focus Loss time distribution (i.e., time that participants spend clicking outside the task browser page during the assessment) and within the top 10% quantile of the accuracy scores were flagged as possibly cheating.

### 2.6 Usability

Telephone and email helplines were set up to support participants with any technical difficulties. We captured unprompted feedback in a detailed log. Qualitative data comprising correspondence between NSHD personnel and study members were examined using thematic analysis to provide insights into the study members’ experiences [26].

Correspondences were anonymised and compiled into NVivo [27, 28], a software for qualitative and mixed methods research. The transcripts of the email conversations consisted of 200 emails (9475 words). Detailed notes were taken by the NSHD personnel when responding to telephone queries, consisting of 78 phone calls (2306 words). Themes from the data were generated using the revised steps suggested by Braun and Clarke (2019) [26]. All steps were completed by one rater. The same participants could contact the helpline with multiple queries, which could result in several thematic codes.

## 3. RESULTS

### 3.1 Sample Features

Out of the 1776 members contacted, 1010 (56.9%) consented to participate, and 813 consented individuals attempted the battery (80.5%). Participants who attempted the battery had an almost balanced female-to-male ratio (51.4% female), predominantly achieved their highest education at A-level (34.6%) and were mostly right-handed (92.4%). In addition, since they were a British birth cohort, they were all 77-year-old White participants with English as a first language. Distributions of demographic factors, socioeconomic status and health-related features are presented in Supplementary Table 3 and Supplementary Figures 1-3.

### 3.2 Adherence

#### 3.2.1 Completion rate for the battery and tasks

Among the 813 participants who attempted the battery, 722 (88.8%) completed all 13 tasks in the assessment (Figure 3A). The average completion time was 40.68 ± 4.24 minutes (Table 1).

**Figure 3.**
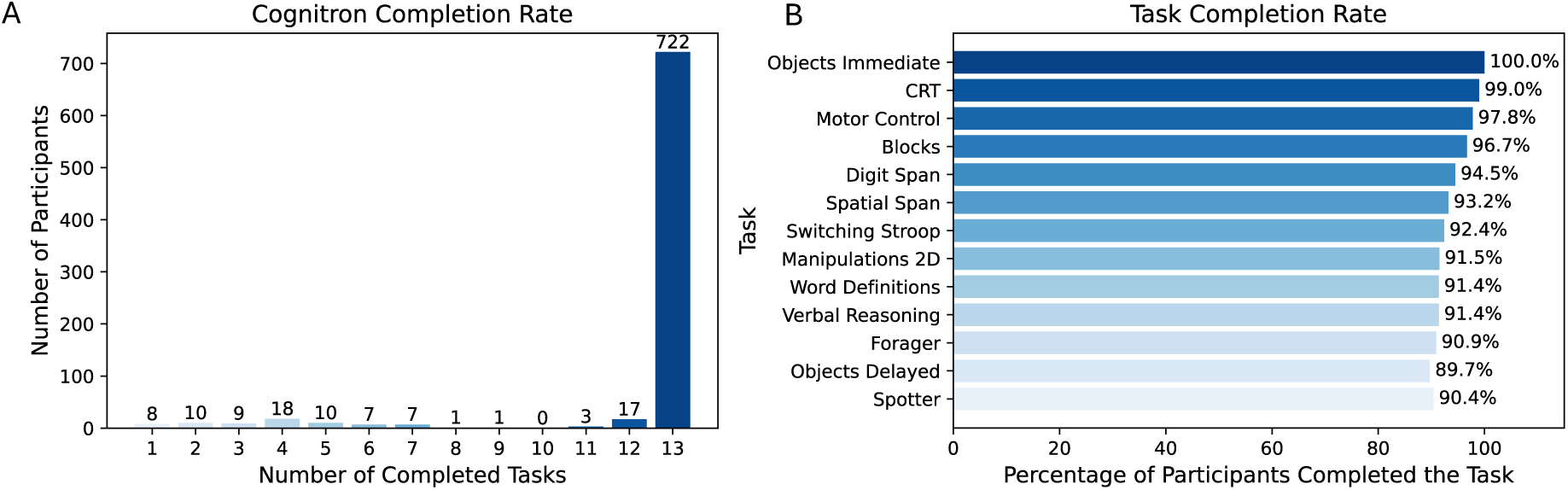
Completion rate for the battery and tasks. A) Cognitron completion rate, indicating the number of participants with various counts of completed tasks. B) Task completion rate, with tasks arranged in order from top to bottom according to the battery sequence.

**Table 1.**
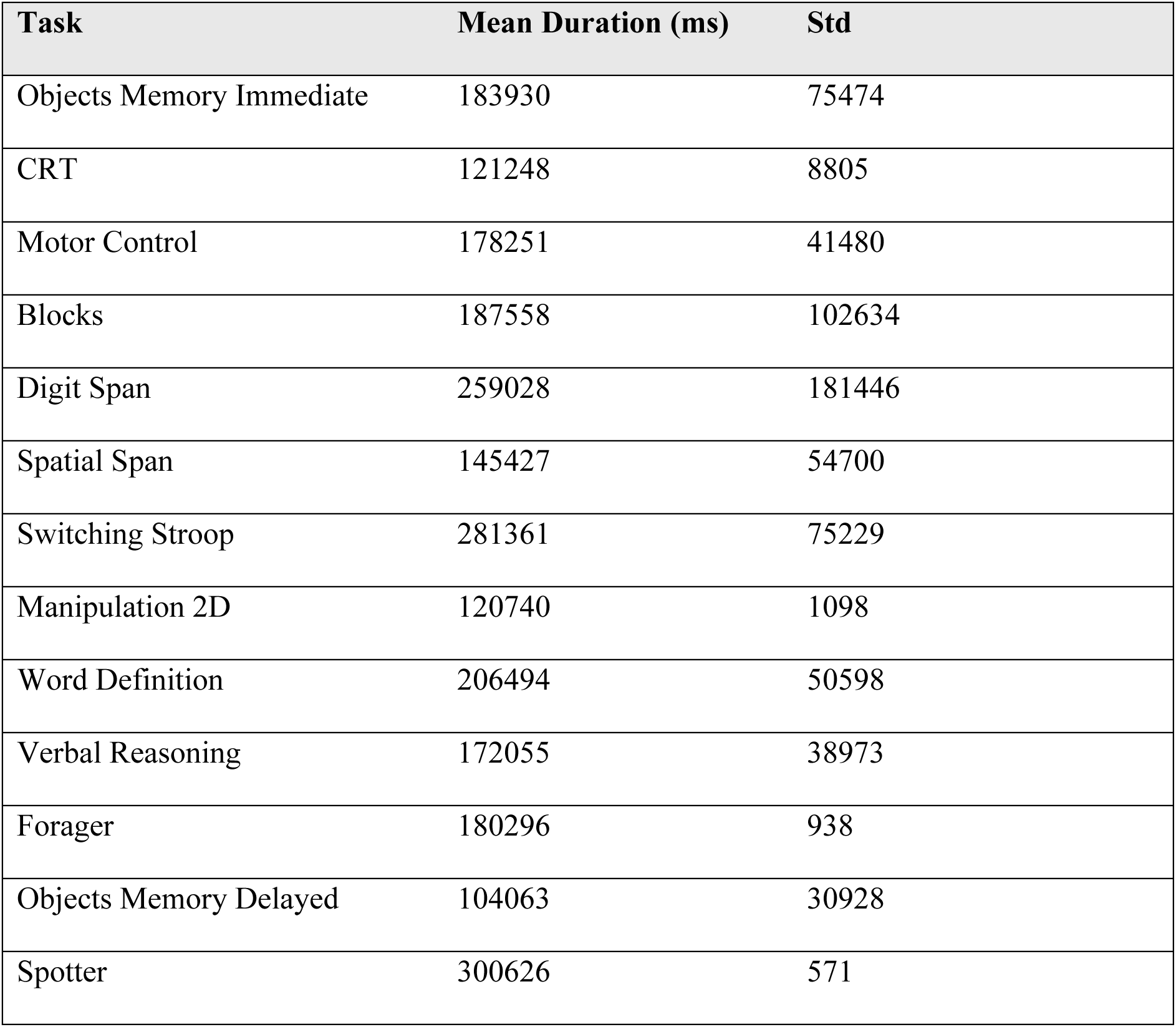
Mean duration in milliseconds for each task.

Most of the participants with 12 tasks completed all but the Objects Delayed (N=10), which was expected because it is not administered if it is not completed within a fixed timeframe on the same day as the Objects Immediate task. More details on the completion rate are in Figure 3B. As expected, Objects Immediate (the first task) had a 100% completion rate. The last task (Spotter) had a completion rate of 90.4%.

#### 3.2.2 Most tasks were characterised by a 100% compliance rate

The results of the task compliance and cheating analysis are reported in Figure 4 and Table 2. Of the 13 tasks studied, eight had a compliance rate of 100%. In the other 4 tasks (Spatial Span, Digit Span, 2D Manipulations and Spotter), less than 2% of participants showed signs of possible non-compliance. The most common sign of non-compliance in these tasks consisted in participants having an accuracy score lower than the expected threshold (e.g., failing the simplest trials).

**Figure 4.**
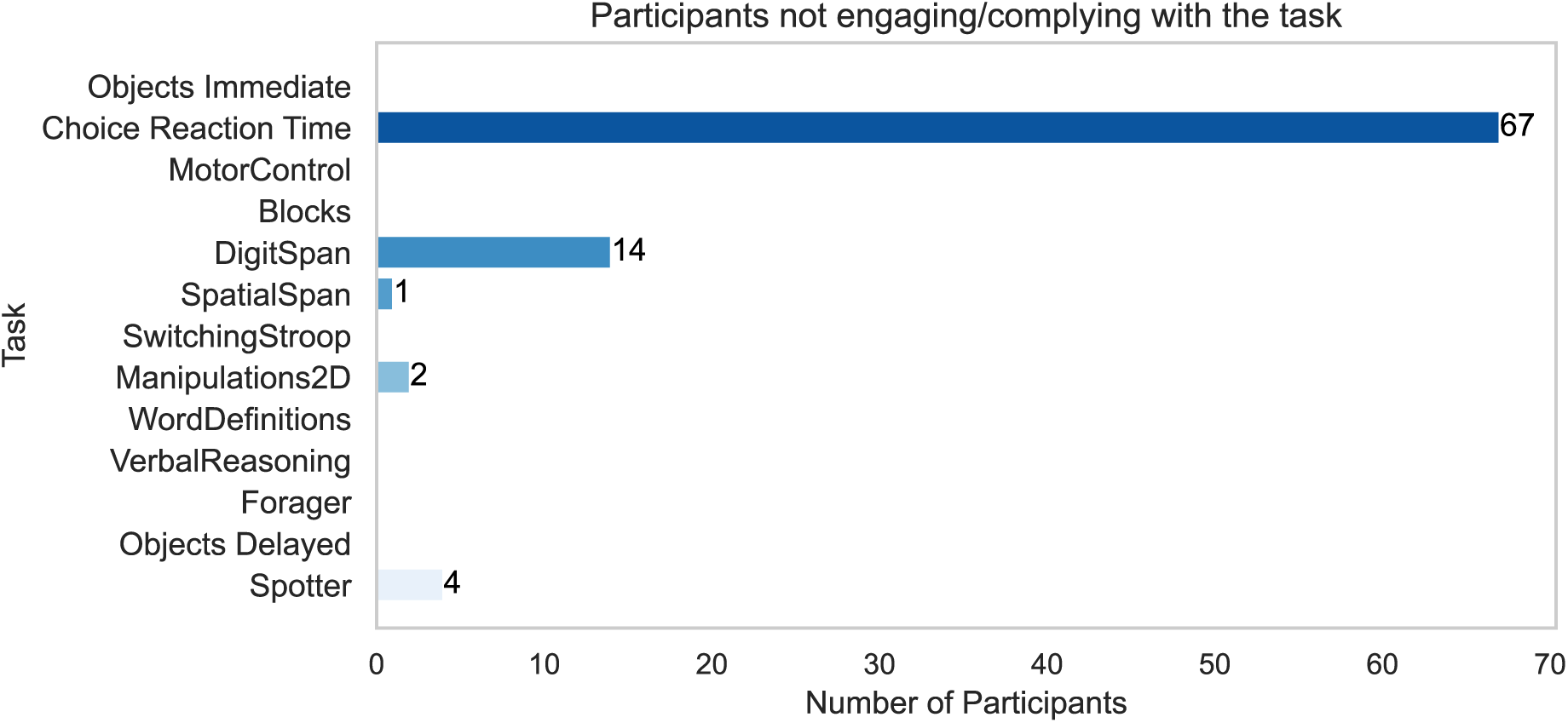
Non-compliance in tasks. The number of participants who did not comply with the cognitive tasks included in the assessment. The higher non-compliance rate for CRT was due to a software error.

**Table 2.**
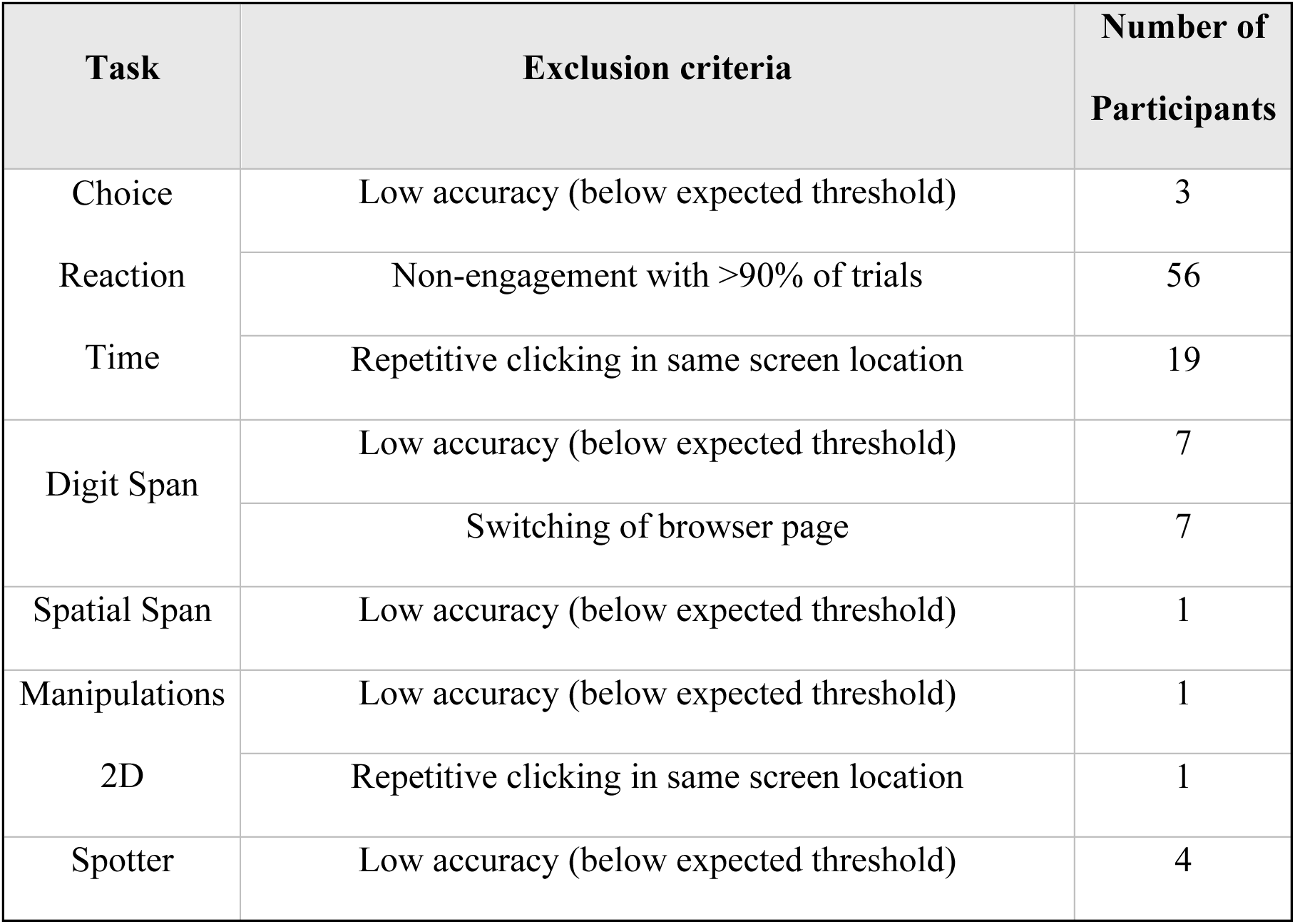
Number of participants who met exclusion criteria for each cognitive task included in the assessment.

Specifically, CRT was characterised by 9% (N=67) of participants who had issues such as not recording more than 90% of the trials (N=56) or clicking repetitively in the same screen location (N=19). The issues were mainly due to task design problems rather than participant compliance. Detailed numbers in Table 2.

### 3.3 Uptake

#### 3.3.1 Higher education, greater adult verbal memory, and regular alcohol consumption correlate with an increased likelihood of study consent

The multivariable logistic regression model was significant in predicting study consent (pseudo R^2^=0.09, N=1137, LLR p < 0.001) (Supplementary Table 4). According to the Wald test, education level (χ²=9.70, p=0.046), word list learning test at age 69 (χ²=11.8, p=0.003) and alcohol consumption (χ²=8.6, p=0.035) were significant predictors. Specifically, compared to members without any educational attainment, individuals with a degree were more likely to consent (p=0.01). Regarding the word list learning test, those who scored at 10% top (p=0.001) and 80% middle were more likely to consent to the study (p=0.003) than those with the lowest 10% score. Finally, members with a higher level of alcohol consumption were more likely to consent to the study (‘four or more times a week’, p=0.012, and ‘two or three time per week’, p=0.047, Supplementary Table 4, Figure 5A).

**Figure 5.**
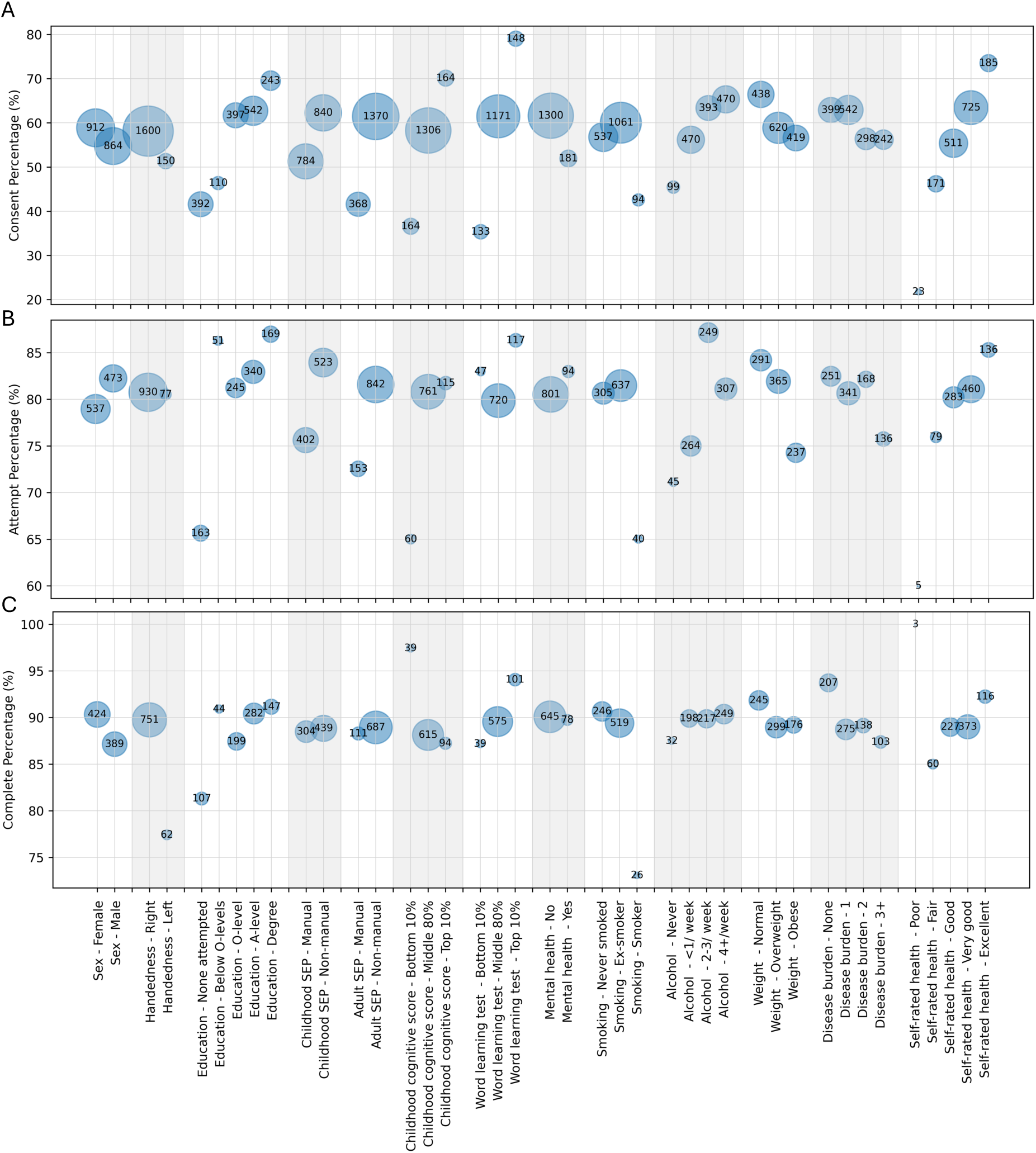
Relationship between uptake of online cognitive tasks and sociodemographic and health-related features. The size of each circle is proportional to the number of participants in the corresponding category, with the numbers also displayed within the circles. The position of the circles represents the percentage of participants in each category who consented, attempted or completed the online battery in panels A, B, and C, respectively.

#### 3.3.2 Participants with moderate alcohol consumption and healthier weights are more likely to attempt the battery

The multivariable logistic regression model that predicts battery attempt was significant (psuedo R^2^ =0.08, N=724, LLR p < 0.05) (Supplementary Table 4). The Wald test identified alcohol consumption (χ²=10.3, p=0.02) and weights (χ²=6.3, p=0.04) as significant predictors. Members with a higher level of alcohol consumption were more likely to attempt the battery (‘4 or more times a week’ p=0.01, ‘two or three time per week’ p=0.002), compared to non-drinkers. Regarding weights, obese members were less likely to attempt the battery (p=0.04, Supplementary Table 4, Figure 5B).

#### 3.3.3 Battery completion is less likely among male, left-handed, current smokers, and individuals with higher disease burden and lower performance in the first task

The multivariable logistic regression model predicting battery completion was significant (pseudo-R²=0.01, N=585, LLR p=0.04) (Supplementary Table 4). The Wald test identified four significant predictors of battery completion: sex (χ²=4.67, p=0.03), handedness (χ²=4.44, p=0.04), smoking by age 69 (χ²=6.07, p=0.048), and disease burden at age 69 (χ²=8.32, p=0.04). Specifically, male participants (p=0.03), left-handed participants (p=0.04), and current smokers (p=0.02) were less likely to complete the battery, respectively. Additionally, an increased disease burden was associated with a lower likelihood of completing the battery (disease burden of 1, 2, 3+, has p value of 0.01, 0.04 and 0.01, respectively). The rates of completing the whole battery for participants with no disease burden, and those with 1, 2, and 3+ disease burdens were 93.7%, 88.3%, 89.1%, and 86.4%, respectively (Figure 5C). Due to the low number of observations, a sensitivity analysis was conducted for battery completion. Specifically, univariate regression models were fitted for individual variables, resulting in OR within a similar range (Supplementary Table 5).

Furthermore, performance in the first cognitive task was also a significant predictor of battery completion (N=813, LLR p=0.03), with higher summary scores correlating with a greater likelihood of completion (p=0.03; Supplementary Table 6).

### 3.4 Usability

#### 3.4.1 Thematic analysis (TA) of qualitative feedback

The inductive TA (Braun & Clarke, 2006) identified five key themes (Figure 6, Supplementary Table 7) summarising the reasons participants contacted the helpline: technical issues (N=137), general queries (N=72), reasons for withdrawing (N=72), subjective reports (N=44) and providing feedback (N=43).

**Figure 6.**
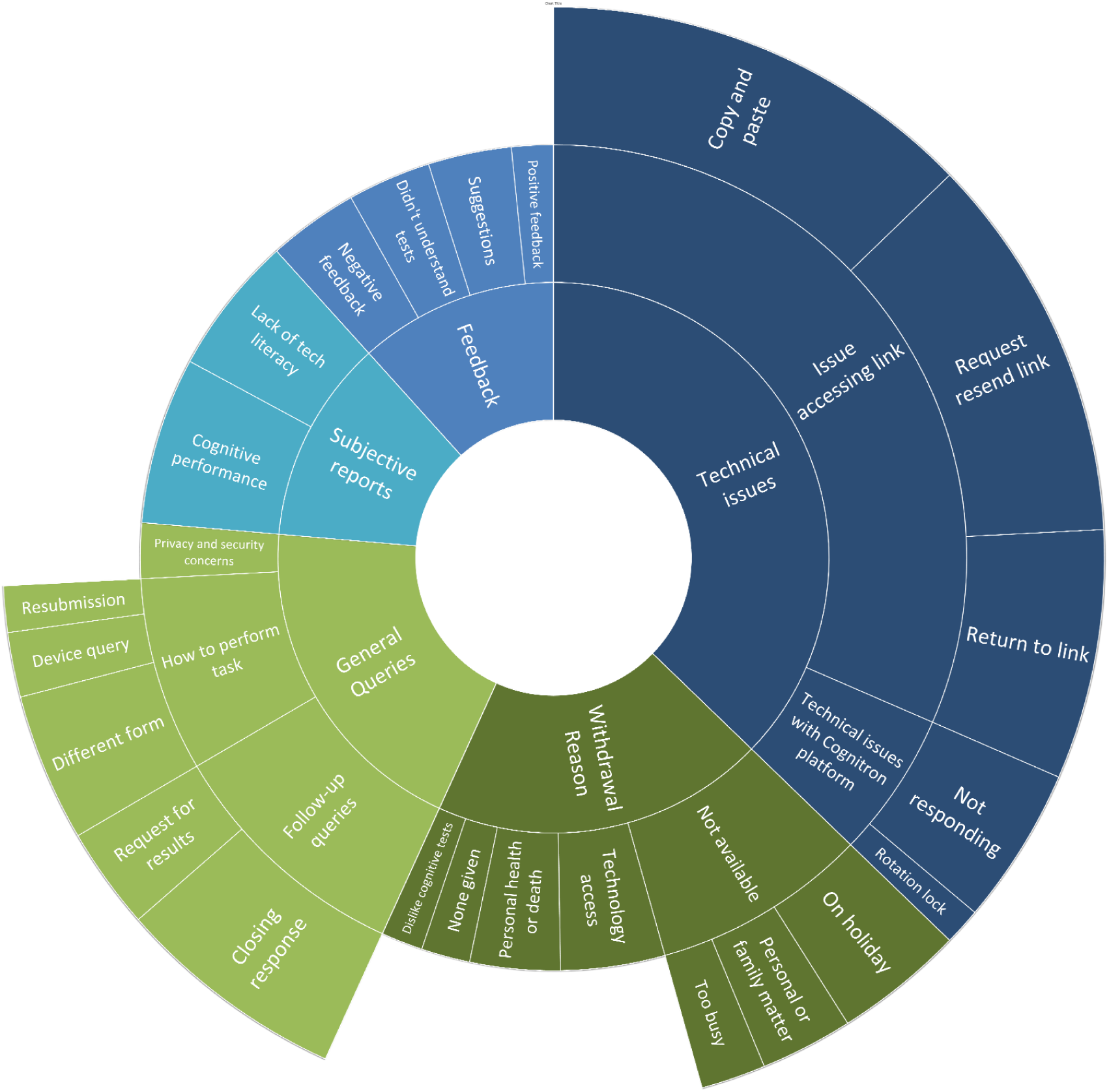
Qualitative feedback for online cognitive tasks. The sunburst diagram represents the results of the qualitative analysis, showing the proportion of data for each theme (inner circle) and subtheme.

#### 3.4.2 Technical issues

The technical issues were divided into two subthemes: issue accessing the link (N=116) and issues with the platform (N=21).

Participants were sometimes unable to navigate between the Qualtrics consent website and the Cognitron platform. After consenting, participants received their user-specific link and were instructed to either copy and paste the link into another window or click the emailed link. This subtheme could be further divided into three issues: unable to ‘copy and paste’ (N=47), ‘requests to resend link’ (N=42) and unable to ‘return to link’ (N=27) after an hour of inactivity. Fewer participants contacted the helpline for difficulties with the Cognitron website. Enquiries were categorized into two subthemes: ‘Cognitron website not responding’ (N=17) and ‘issues with rotation lock on tablet device’ (N=4). The webpage did not respond due to poor internet connection or outdated web browsers. The rotation lock occurred on tablets as the Cognitron platform requests portrait screen orientation.

#### 3.4.3 General Queries

General queries had three subthemes: follow-up queries (N=36), how to perform the task (N=28) and privacy and security concerns (N=8). These subthemes included participants who made a ‘request for results’ (N=11), while others provided a closing response (N=25) (e.g., “I have now completed the tasks”). Participants who asked ‘how to perform the task’ either had a ‘device query’ (N=7), asked if they could complete the tasks in a ‘different form’ (N=16) or asked if they could go back to the tasks later - ‘resubmission’ (N=5). Some participants (N=8) had privacy and security concerns (e.g., “What happens between it [the data] being sent and received?”).

#### 3.4.4 Reasons for withdrawing from Online46

The withdrawing reasons were dislike of cognitive tasks (N=6), none given (N=7), not available (N=31), personal health or death (N=13) and technology access (N=15). Study members were not available for three reasons: on holiday (N=14), personal or family matters (N=10) and too busy (N=7).

#### 3.4.5 Subjective reports

The subjective reports provided insights into participants’ perceptions of online cognitive testing and can be divided into two subthemes: lack of technical literacy (N=20) and cognitive performance (N=24). The lack of technical literacy subtheme highlights the participants’ perception of “modern on-line technology”. Eight participants also provided rich commentary on their perceived cognitive performance. This personal commentary highlighted the participants’ negative perception of their cognition.

#### 3.4.6 Providing feedback

The helpline received negative (N=13) and positive feedback (N=6), suggestions (N=12) and comments from those who did not understand the tasks (N=12).

## 4. DISCUSSION

We completed a quantitative and qualitative analysis of the uptake, adherence and usability of unsupervised computerized cognitive assessments in an elderly general population birth cohort, with the goal of informing future studies and paving the way to the application of such assessments in clinical research and practice.

We confirmed that the participation rate of elderly cohorts at different stages of recruitment is adequate. Specifically, the overall consent rate of 56.9%, of whom 80.5% attempted and 88.8% completed the battery, were comparable to previous sub-studies conducted under supervised conditions [20]. At just above 55%, the consent rate was similar to Insight 46 (a deep-phenotyping sub-study of the NSHD), where out of the 1322 eligible participants, around 59% were willing to attend a clinic visit in London, and out of the 841 members invited 60% were recruited and attended a research visit [20].

The duration of the assessment (40.68 ± 4.24 minutes) was within the range of other self-administrated online cognitive assessments such as the Touch panel-type dementia assessment scale (30 minutes) [29], and short-form MicroCog (30-45 minutes). The qualitative feedback did not indicate any complaints for the duration of the assessment [30, 31]; therefore, a cognitive assessment of around 40 minutes may be well received by elderly cohorts, although shorter batteries are likely to be more appropriate when repeat testing over short periods is required.

Our study showed that consent, participation and completion of the online assessment related to sociodemographic and health-related factors. Higher education increases the likelihood of consenting to the assessment, likely due to greater willingness to learn and use technology [32], improved understanding of research information [33], and increased confidence in performance [34, 35]. This aligns with previous findings from NSHD sub-studies (i.e., Insight46) [20, 34], other web-based cognitive batteries [36] and epidemiological studies [37–39]. Similarly, participants with higher past cognitive scores were more likely to consent to the battery, which could be explained by an overall greater health literacy [34, 40, 41].

Interestingly, participants who consume alcohol more than twice a week were more likely to consent and attempt the battery compared to non-drinkers. Previous studies identified an association between moderate alcohol consumption and better education, higher socioeconomic status [42, 43] and better psychological and physical well-being [44, 45], which could make regular and moderate drinkers more proactive in the participation in health assessments. Supporting this hypothesis, participants who completed all tasks or had higher completion rates were less likely to be current smokers, obese, or have a high disease burden. This suggests that individuals with better health are more likely to complete the battery.

Overall, these biases should be considered when interpreting results on this cohort, as the strength of associations might not be representative of the whole population; however, this bias might not be problematic in the case of studies assessing the relationship between health and exposures across the life course [46]. This was also the case for previous NSHD studies [17, 20].

The qualitative feedback allowed us to identify recommendations for improving accessibility in online cognitive assessment of elderly cohorts (Table 3). First, although compliance rates for individual tasks were generally very high, the process of articulating from recruitment through to starting the battery should be streamlined to prevent challenges and dropout when moving between platforms and task stages. Clear concise written or video instructions should be provided before each stage of the battery and the inclusion of practice trials, which may not be needed in younger participants, is advisable. Large fonts and/or audio instructions are important for elderly cohorts and user-friendly interfaces with clear language and intuitive icons. According to the qualitative feedback, participants differ in whether they want to see reports of their performance. A simple interpretable summary of performance should be provided at the end of the assessment but shown only to participants who opt to see it. Then, a detailed Q&A should be available with task instructions and a summary of privacy and data sharing regulations. This should be complementary to an active helpline to address any potential engagement and technical issue. Finally, most elderly participants who begin an online assessment tend to finish it – the greatest bias occurs at the point of recruitment into the study. Further investigation is warranted into how to increase engagement at the recruitment stage.

**Table 3.** List of challenges and recommendations identified through the qualitative analysis.

|  | Recommendations |
| --- | --- |
| Stressful tests | Opportunity to complete the assessment across multiple days, with possibility to easily re-access the platform where the test is hosted and system of reminders |
| Issues accessing the tests online | 1. Detailed video / written instructions to guide participants on how to access the assessment<br>2. Simplification of the process by having the consent and assessment within the same platform |
| Feedback and results from online assessment | 1. Design of automatic results' reports with summary of cognitive abilities<br>2. Possibility for users to decide whether to see the results or not, to prevent negative feelings towards their cognitive abilities<br>3. Possibility for users to obtain either a summary of performance or a measure of comparison towards other participants |
| Issues with test website freezing or not responding | 1. Reminder to ensure testing environment has stable internet connection<br>2. Availability of phone and / or email helpline to handle potential issues with the assessment platform |
| Older adults physical condition interrupts or hinders the completion of assessment | 1. Design assessments to allow pausing between tests and restarting it on the same day (or another day)<br>2. Design of the interface such that it is easier for older cohorts (e.g., bigger font sizes, possibility to hear written instructions, limited flashing lights ...) |
| Lack of technical literacy | 1. Design of simplified interface with clear language and intuitive icons<br>2. Simplification of the consent and assessment process |
| Uncertainty for how to conduct the test | 1. Detailed Q&A webpage<br>2. Task-specific instructions before each task<br>3. Practice trials before the tests<br>4. Interactive tutorials that walk participants through the practice sessions |
| Unavailability to complete the test | 1. Possibility to complete the assessment throughout a longer timespan<br>2. Multiple follow-up emails |
| Privacy and authenticity concerns | 1. Use of previously known email domain when contacting participants<br>2. Advertisement of study in email and / or paper newsletters<br>3. Easily accessible summary of privacy and data-sharing regulations (e.g., inclusion in the Q&A webpage) that is not embedded in extensive text |

Our study has some limitations. First, quantifying factors that underlie self-selection bias was a focus of the study; however the sample is a birth British cohort, which means that variability in response and compliance with age and ethnicity could not be evaluated. Some sub-groups were too small to evaluate reliably, e.g., those who were left-handed showed lower completion rates here, but such bias has not been evident in other older adult [12] larger population studies using the same technology [22]. Additionally, out of the 2360 active NSHD members, 583 individuals did not have email or internet access and were not included in the uptake analysis. Feedback was collected from participants with issues while conducting the assessment, which tends to highlight negative not positive aspects of online assessments; the high completion rates support that in general the assessment was well received. Information about participants’ retention and attrition rates with repeat online assessment will only be available when future data are collected. Further research will explore the association between online cognitive assessment results and biomarkers, with findings to be published separately. Given the adequate participation rate and the demographic predictors identified for an elderly cohort, this online cognitive battery is not only valuable for neuroscience studies but also has broad applicability in fields such as demography, economics, and education research.

In conclusion, online cognitive assessments are feasible for elderly cohorts if design considerations pertaining to accessibility, engagement and compliance are followed.

## Data Availability

The data analysed in the present study are available via the NSHD data access process(https://nshd.mrc.ac.uk/data-sharing/)

## FUNDING

MR, MP, AW are funded by the Medical Research Council (MC_UU_00019/1 and 3). VG is supported by the Medical Research Council, MR/W00710X/1. MDG is funded by Imperial College London. ZC is funded by Kings College London. Insight 46 is funded by grants from Alzheimer’s Research UK (ARUK-PG2014-1946, ARUK-PG2017-1946), Alzheimer’s Association (SG-666374-UK BIRTH COHORT), the Medical Research Council Dementias Platform UK (CSUB19166), The Wolfson Foundation (PR/ylr/18575), The Medical Research Council (MC_UU_10019/1, MC_UU_10019/3), and Brain Research Trust (UCC14191). JMS J.M.S. is supported by University College London Hospitals Biomedical Research Centre.

## CONFLICT OF INTEREST STATEMENT

The funders of the study had no role in study design, data collection, analysis, interpretation, report writing, or in the decision to submit the article for publication. AH is owner/director of H2 Cognitive Designs Ltd and Future Cognition Ltd, which produce online assessment technology and provide online survey data collection for third parties. PH is founder and director of H2 Cognitive Designs LTD, which develops and markets online cognitive tasks.

## CONSENT STATEMENT

The study was approved by the National Research Ethics Service Committee London (REC reference 14/LO/1173) and all participants provided written informed consent.

## Notes

### Author Declarations

National Research Ethics Service Committee London (REC reference 14/LO/1173) gave ethical approval for this work

### Summary of Updates

Author information updated: change of order

## REFERENCES

1. Nasreddine, Z.S., et al., The Montreal Cognitive Assessment, MoCA: a brief screening tool for mild cognitive impairment. J Am Geriatr Soc, 2005. 53(4): p. 695–9.

2. Del Giovane, M., et al., Computerised cognitive assessment in patients with traumatic brain injury: an observational study of feasibility and sensitivity relative to established clinical scales. EClinicalMedicine, 2023. 59: p. 101980.

3. Brooker, H., et al., FLAME: A computerized neuropsychological composite for trials in early dementia. Alzheimers Dement (Amst), 2020. 12(1): p. e12098.

4. Giunchiglia, V., et al., Neural correlates of cognitive ability and visuo-motor speed: Validation of IDoCT on UK Biobank Data. Imaging Neuroscience, 2024. 2: p. 1–25.

5. Gruia, D.-C., et al., Online monitoring technology for deep phenotyping of cognitive impairment after stroke. medRxiv, 2024: p. 2024.09.06.24313173.

6. Koo, B.M. and L.M. Vizer, Mobile Technology for Cognitive Assessment of Older Adults: A Scoping Review. Innov Aging, 2019. 3(1): p. igy038.

7. Vaportzis, E., M.G. Clausen, and A.J. Gow, Older Adults Perceptions of Technology and Barriers to Interacting with Tablet Computers: A Focus Group Study. Front Psychol, 2017. 8: p. 1687.

8. Lawry, S., et al., Age, familiarity, and intuitive use: An empirical investigation. Applied Ergonomics, 2019. 74: p. 74–84.

9. Wilson, J., et al., Barriers and facilitators to the use of e-health by older adults: a scoping review. BMC Public Health, 2021. 21(1): p. 1556.

10. Corbett, A., et al., Cognitive decline in older adults in the UK during and after the COVID-19 pandemic: a longitudinal analysis of PROTECT study data. The Lancet Healthy Longevity, 2023. 4(11): p. e591–e599.

11. Shibata, K., et al., Remote digital cognitive assessment reveals cognitive deficits related to hippocampal atrophy in autoimmune limbic encephalitis: a cross-sectional validation study. eClinicalMedicine, 2024. 69: p. 102437.

12. Bălăeţ, M., et al., Online cognitive monitoring technology for people with Parkinson’s disease and REM sleep behavioural disorder. npj Digital Medicine, 2024. 7(1): p. 118.

13. Feenstra, H.E.M., et al., Reliability and validity of a self-administered tool for online neuropsychological testing: The Amsterdam Cognition Scan. J Clin Exp Neuropsychol, 2018. 40(3): p. 253–273.

14. Cyr, A.A., K. Romero, and L. Galin-Corini, Web-Based Cognitive Testing of Older Adults in Person Versus at Home: Within-Subjects Comparison Study. JMIR Aging, 2021. 4(1): p. e23384.

15. Sokołowski, D.R., et al., Participation and engagement in online cognitive testing. Scientific Reports, 2024. 14(1): p. 14800.

16. Kuh, D., et al., Cohort profile: updating the cohort profile for the MRC National Survey of Health and Development: a new clinic-based data collection for ageing research. Int J Epidemiol, 2011. 40(1): p. e1–9.

17. Kuh, D., et al., The MRC National Survey of Health and Development reaches age 70: maintaining participation at older ages in a birth cohort study. European Journal of Epidemiology, 2016. 31(11): p. 1135–1147.

18. Lane, C.A., et al., Study protocol: Insight 46 – a neuroscience sub-study of the MRC National Survey of Health and Development. BMC Neurology, 2017. 17(1): p. 75.

19. Murray-Smith, H., et al., Updating the study protocol: Insight 46 - a longitudinal neuroscience sub-study of the MRC National Survey of Health and Development - phases 2 and 3. BMC Neurol, 2024. 24(1): p. 40.

20. James, S.N., et al., Using a birth cohort to study brain health and preclinical dementia: recruitment and participation rates in Insight 46. BMC Res Notes, 2018. 11(1): p. 885.

21. Moulton, V., McElroy, E., Richards, M., Fitzsimons, E., Northstone, K., Conti, G., Ploubidis,G.B., Sullivan, A., O’Neill, D., A guide to the cognitive measures in five British birth cohort studies. London, UK: CLOSER. 2020.

22. Hampshire, A., et al., Cognition and Memory after Covid-19 in a Large Community Sample. N Engl J Med, 2024. 390(9): p. 806–818.

23. Giovane, M.D., et al., Computerised cognitive testing and multi-domain structural Magnetic Resonance Imaging in patients with idiopathic Normal Pressure Hydrocephalus and Alzheimer’s disease. Alzheimer’s & Dementia, 2023. 19(S18): p. e074654.

24. Cheetham, N.J., et al., The effects of COVID-19 on cognitive performance in a community-based cohort: a COVID symptom study biobank prospective cohort study. eClinicalMedicine, 2023. 62: p. 102086.

25. Valentina Giunchiglia, D.G., Annalaura Lerede, William Trender, Peter Hellyer, Adam Hampshire, Iterative decomposition of visuomotor, device and cognitive variance in large scale online cognitive test data. Research Square, 2023.

26. Braun, V. and V. Clarke, Reflecting on reflexive thematic analysis. Qualitative Research in Sport, Exercise and Health, 2019. 11(4): p. 589–597.

27. Woods, M., et al., Advancing Qualitative Research Using Qualitative Data Analysis Software (QDAS)? Reviewing Potential Versus Practice in Published Studies using ATLAS.ti and NVivo, 1994–2013. Social Science Computer Review, 2015. 34(5): p. 597–617.

28. Zamawe, F.C., The Implication of Using NVivo Software in Qualitative Data Analysis: Evidence-Based Reflections. Malawi Med J, 2015. 27(1): p. 13–5.

29. Inoue, M., et al., Touch Panel-type Dementia Assessment Scale: a new computer-based rating scale for Alzheimer’s disease. Psychogeriatrics, 2011. 11(1): p. 28–33.

30. Shermon, E., L.O. Vernon, and A.J. McGrath, Cognitive assessment of elderly inpatients: a clinical audit. Dement Geriatr Cogn Dis Extra, 2015. 5(1): p. 25–31.

31. Lynham, A.J., I.R. Jones, and J.T.R. Walters, Cardiff Online Cognitive Assessment in a National Sample: Cross-Sectional Web-Based Study. J Med Internet Res, 2023. 25: p. e46675.

32. Bailey, C. and C. Sheehan, Technology, older persons’ perspectives and the anthropological ethnographic lens. Alter, 2009. 3(2): p. 96–109.

33. Asare, M., M. Flannery, and C. Kamen, Social Determinants of Health: A Framework for Studying Cancer Health Disparities and Minority Participation in Research. Oncol Nurs Forum, 2017. 44(1): p. 20–23.

34. Stafford, M., et al., Using a birth cohort to study ageing: representativeness and response rates in the National Survey of Health and Development. Eur J Ageing, 2013. 10(2): p. 145–157.

35. Chatfield, M.D., C.E. Brayne, and F.E. Matthews, A systematic literature review of attrition between waves in longitudinal studies in the elderly shows a consistent pattern of dropout between differing studies. J Clin Epidemiol, 2005. 58(1): p. 13–9.

36. Lynham, A.J., I.R. Jones, and J.T.R. Walters, Web-Based Cognitive Testing in Psychiatric Research: Validation and Usability Study. J Med Internet Res, 2022. 24(2): p. e28233.

37. Galea, S. and M. Tracy, Participation Rates in Epidemiologic Studies. Annals of Epidemiology, 2007. 17(9): p. 643–653.

38. Gallicchio, L., et al., Utilizing SEER Cancer Registries for Population-Based Cancer Survivor Epidemiologic Studies: A Feasibility Study. Cancer Epidemiol Biomarkers Prev, 2020. 29(9): p. 1699–1709.

39. Nikolaou, E., et al., Assessing the use of minimally invasive self-sampling at home for long-term monitoring of the microbiota within UK families. Scientific Reports, 2023. 13(1): p. 18201.

40. Rydén, L., et al., Attrition in the Gothenburg H70 birth cohort studies, an 18-year follow-up of the 1930 cohort. Front Epidemiol, 2023. 3: p. 1151519.

41. Federman, A.D., et al., Health literacy and cognitive performance in older adults. J Am Geriatr Soc, 2009. 57(8): p. 1475–80.

42. Collins, S.E., Associations Between Socioeconomic Factors and Alcohol Outcomes. Alcohol Res, 2016. 38(1): p. 83–94.

43. Dunbar, R.I.M., et al., Functional Benefits of (Modest) Alcohol Consumption. Adaptive Human Behavior and Physiology, 2017. 3(2): p. 118–133.

44. Grønkjær, M., et al., Prospective associations between alcohol consumption and psychological well-being in midlife. BMC Public Health, 2022. 22(1): p. 204.

45. Towers, A., et al., The “Health Benefits” of Moderate Drinking in Older Adults may be Better Explained by Socioeconomic Status. The Journals of Gerontology: Series B, 2016. 73(4): p. 649–654.

46. Rothman, K.J., J.E. Gallacher, and E.E. Hatch, Why representativeness should be avoided. International Journal of Epidemiology, 2013. 42(4): p. 1012–1014.

