## Supplementary Files for "Online46: online cognitive assessments in elderly cohorts - the British 1946 birth cohort case study"

**Supplementary Table 1 - Cognitive task designs.** The tasks involved in the study were Object Memory Immediate, Choice Reaction Time, Precision Motor Control, Blocks, Digit Span, Spatial Span, Switching Stroop, Manipulations 2D, Word Definition, Verbal Reasoning, Forager, Object delay and Spotter.

| **Task** | **Designs** |
| --- | --- |
| Objects Memory | Participants are presented with images of target objects. They must identify the target object from a set of variations of the original image immediately at the end of task (Objects Immediate) or after a delay (Objects Delayed). |
| Choice Reaction Time (CRT) | Participants are presented with an arrow pointing either left or right and must respond by clicking on the same side of the screen where the arrow is directed. |
| Motor Control | Participants are presented with a moving red target that appears in unpredicted locations on the screen and must click on it quickly and accurately. |
| Block | Participants are shown a grid with colored blocks and a grid with grey blocks. They need to remove a specified number of blocks from the colored grid to achieve the target configuration shown in a grey grid. |
| Digit Span | Participants are shown a sequence of digits to memorise and recall. Each correct trial results in one more digit in the sequence for the next trial. The task ends after three consecutive incorrect trials. |
| Spatial Span | Participants are presented with a series of grey squares displayed at different positions on a 4x4 grid and are asked to memorise and recall the sequence. Each correct trial results in one more square in the sequence for the next trial. The task ends after three consecutive incorrect trials. |
| Switching Stroop | Participants are presented with a box in either blue or red color, with the words 'BLUE' and 'RED' written in matching or non-matching ink colors on the two sides of the central box. If the instruction displayed at the top of the box is 'INK,' participants need to click the word with ink color same as the box color; if the instruction is 'TEXT,' participants select the word describing the box color. |
| Manipulation 2D | Participants are presented with a target configuration—a grid with colored squares positioned at different locations. Participants then need to identify a grid from four probes that can match the target configuration through rotation. |
| Word Definitions | Participants are presented with English word and are required to choose its corresponding definition from four options within a set time limit. |
| Verbal Reasoning | Participants are presented with various combinations of geometric shapes and need to determine if the statement describing their relative position is true or false. |
| Forager | Participants are shown continuous sequence of shapes including squares, diamonds, triangles, and circles. They need to click on the shapes to determine which one is likely to score points based on the feedback they receive (correct/incorrect). After successfully following the rule for 6 consecutive trials, a new rule will be introduced (i.e., the best shape change) and the participants will receive negative feedback. |
| Spotter | Participants are presented with numbers which appear briefly on the screen within a pixelated square. They are asked to click on the square as soon as they see a “0”. |

**Supplementary Table 2 - Description of sociodemographic factors and health features.** The variables involved in the analysis for the uptake of the online battery were 1) Sex, 2) Handedness, 3) Education, 4) Childhood socioeconomic position (SEP), 5) Adult SEP, 6) Childhood cognitive score, 7) Word learning test memory score at age 69, 8) Mental health prevalence at age 69, 9) Lifetime smoking by age 69, 10) Alcohol use at age 69, 11) Weight status at age 69, 12) Overall disease burden at age 69, and 13) Self-rated health at age 68.

| **Variable** | **Description** |
| --- | --- |
| Sex | Male and Female |
| Handedness | Right and left-handed |
| Education | The highest educational level achieved by age 26. The categories of education included non-attempted, below O level, O level, A-level and degree. |
| Childhood socioeconomic position (SEP) | It was determined based on paternal occupation in 1961 and categorised into manual (skilled manual, semi-skilled and unskilled) and non-manual (professional, intermediate, skilled non-manual) professions. |
| Adult SEP | It was determined based on participants’ own occupation at age 53, the performance was divided into lowest 10%, middle 80% and highest 10%. |
| Childhood cognitive score | It was obtained from tests for verbal and non-verbal ability ^48^. |
| Word learning test memory score at 69 years | The sum of scores (maximum 45) from 3-word list memory tests completed at age 69. The performance was divided into lowest 10%, middle 80% and highest 10%. |
| Mental health prevalence at age 69 | It was derived from 28-item version of the General Health Questionnaire completed at age. |
| Lifetime smoking by 69 | The history of smoking was derived from a questionnaire completed at age 68, or 69 in case of missing data in the previous year, and grouped participants into never, ex and current smoker. |
| Alcohol use at age 69 | Alcohol consumption was noted as: never, less than once a week, 2-3 times per week and 4 times per week. |
| Weight status at age 69 | BMI was calculated based on height and weight recorded in 2015 and categorized into normal (body mass index (BMI) < 25.0 kg/ m2), overweight (BMI of 25.0 to <30.0 kg/m2), or obese (BMI>30.0 kg/m2) |
| Overall disease burden at age 69 | The measure of overall disease burden was based on a 19-point doctor-diagnosed chronic disease scale measured at age 68-70 and categorized into 0,1,2,3 and 3+, where the higher is the score, the higher is the burden |
| Self-rated health at age 68 | Participants self-rated their health as poor, fair, good, very good or excellent. |

**Supplementary Table 3 - Numbers and distribution of sociodemographic factors and health features for different levels of participation.** Percentages were calculated relative to the counts within other categories of the same variable.

| **Variable** |  | **NSHD members** | **Consent** | | **Battery Attempt** | | **Battery Completion** | |
| --- | --- | --- | --- | --- | --- | --- | --- | --- |
|  |  |  | **Yes** | **No** | **Yes** | **No** | **Yes** | **No** |
|  | Max number | 1776 | 1010 | 766 | 813 | 197 | 722 | 91 |
| Sex |  |  |  |  |  |  |  |  |
|  | Female | 912 (51%) | 537 (53%) | 375 (49%) | 424 (52%) | 113 (57%) | 383 (53%) | 50 (55%) |
| Handedness | |  |  |  |  |  |  |  |
|  | Right | 1600 (90%) | 930 (92%) | 670 (89%) | 751 (92%) | 179 (92%) | 674 (93%) | 77 (85%) |
| Education | |  |  |  |  |  |  |  |
|  | None attempted | 392 (23%) | 163 (17%) | 229 (32%) | 107 (14%) | 56 (30%) | 87 (13%) | 20 (22%) |
|  | Below O-levels | 110 (7%) | 51 (5%) | 59 (8%) | 44 (6%) | 7 (4%) | 40 (6%) | 4 (4%) |
|  | O-level | 397 (24%) | 245 (25%) | 152 (21%) | 199 (26%) | 46 (24%) | 174 (25%) | 25 (28%) |
|  | A-level | 542 (32%) | 340 (35%) | 202 (28%) | 282 (36%) | 58 (31%) | 255 (37%) | 27 (30%) |
|  | Degree | 243 (14%) | 169 (18%) | 74 (10%) | 147 (19%) | 22 (12%) | 134 (19%) | 13 (15%) |
| Childhood SEP | |  |  |  |  |  |  |  |
|  | Manual | 784 (48%) | 402 (43%) | 382 (55%) | 304 (41%) | 98 (54%) | 269 (41%) | 35 (42%) |
| Adult SEP | |  |  |  |  |  |  |  |
|  | Manual | 368 (21%) | 153 (15%) | 215 (30%) | 111 (14%) | 42 (21%) | 98 (14%) | 13 (15%) |
| Childhood cognitive score | |  |  |  |  |  |  |  |
|  | Bottom 10% | 164 (10%) | 60 (6%) | 104 (15%) | 39 (5%) | 21 (11%) | 38 (6%) | 1 (1%) |
|  | Middle 80% | 1306 (80%) | 761 (81%) | 545 (78%) | 615 (82%) | 146 (78%) | 542 (82%) | 73 (85%) |
|  | Top 10% | 164 (10%) | 115 (12%) | 49 (7%) | 94 (13%) | 21 (11%) | 82 (12%) | 12 (14%) |
| Adult cognitive score | |  |  |  |  |  |  |  |
|  | Bottom 10% | 133 (9%) | 47 (5%) | 86 (15%) | 39 (6%) | 8 (5%) | 34 (5%) | 5 (7%) |
|  | Middle 80% | 1171 (81%) | 720 (81%) | 451 (79%) | 575 (80%) | 145 (86%) | 515 (80%) | 60 (85%) |
|  | Top 10% | 148 (10%) | 117 (13%) | 31 (6%) | 101 (14%) | 16 (9%) | 95 (15%) | 6 (8%) |
| Mental health prevalence at age 69 | | |  |  |  |  |  |  |
|  | Yes | 181 (12%) | 94 (11%) | 87 (15%) | 78 (11%) | 16 (9%) | 70 (11%) | 8 (11%) |
| Lifetime smoking by 69 years | | |  |  |  |  |  |  |
|  | Never smoke | 537 (32%) | 305 (29%) | 232 (33%) | 246 (31%) | 59 (31%) | 223 (32%) | 23 (27%) |
|  | Ex-smoker | 1061(63%) | 637 (71%) | 424 (60%) | 519 (66%) | 118 (62%) | 464 (66%) | 55 (65%) |
|  | Smoker | 94 (5%) | 40 (4%) | 54 (8%) | 26 (3%) | 14 (7%) | 19 (3%) | 7 (8%) |
| Alcohol use at age 69 | |  |  |  |  |  |  |  |
|  | Never | 99 (7%) | 45 (5%) | 54 (10%) | 32 (5%) | 13 (8%) | 28 (4%) | 4 (6%) |
|  | <1 per week | 470 (33%) | 264 (31%) | 206 (36%) | 198 (28%) | 66 (39% | 178 (28%) | 20 (29%) |
|  | 2-3 per week | 393 (27%) | 249 (29%) | 144 (25%) | 217 (31%) | 32 (19%) | 195 (31%) | 22 (31%) |
|  | 4+ per week | 470 (33%) | 307 (35%) | 163 (29%) | 249 (36%) | 58 (34%) | 225 (36%) | 24 (34%) |
| Weight status at age 69 | |  |  |  |  |  |  |  |
|  | Normal | 438 (30%) | 291 (33%) | 147 (25%) | 245 (34%) | 46 (27%) | 225 (35%) | 20 (28%) |
|  | Overweight | 620 (42%) | 365 (41%) | 255 (44%) | 299 (42%) | 66 (38%) | 266 (41%) | 33 (46%) |
|  | Obese | 419 (28%) | 237 (27%) | 182 (31%) | 176 (24%) | 61 (35%) | 157 (24%) | 19 (26%) |
| Overall disease burden at age 69 | | |  |  |  |  |  |  |
|  | None | 399 (27%) | 251 (28%) | 148 (25%) | 207 (29%) | 44 (25%) | 194 (30%) | 13 (18%) |
|  | 1 | 542 (37%) | 341 (38%) | 201 (34%) | 275 (38%) | 66 (38%) | 244 (37%) | 31 (43%) |
|  | 2 | 298 (20%) | 168 (19%) | 130 (22%) | 138 (19%) | 30 (17%) | 123 (19%) | 15 (21%) |
|  | 3+ | 242 (16%) | 136 (15%) | 106 (18%) | 103 (14%) | 33 (19%) | 90 (14%) | 13 (18%) |
| Self-rated health at age 68 | | | |  |  |  |  |  |
|  | Poor | 23 (1%) | 5 (1%) | 18 (3%) | 3 (0.4%) | 2 (1%) | 3 (0.4%) | 0 (0%) |
|  | Fair | 171 (11%) | 79 (8%) | 92 (14%) | 60 (8%) | 19 (10%) | 51 (7%) | 9 (11%) |
|  | Good | 511 (32%) | 283 (29%) | 228 (35%) | 227 (29%) | 56 (30%) | 202 (29%) | 25 (30%) |
|  | Very good | 725 (45%) | 460 (48%) | 265 (41%) | 373 (48%) | 87 (47%) | 332 (48%) | 41 (49%) |
|  | Excellent | 185 (11%) | 136 (14%) | 49 (8%) | 116 (15%) | 20 (11%) | 107 (15%) | 9 (11%) |

SEP=socioeconomic position

**Supplementary Table 4-** **Logistic regression models on demographic variables for battery consent, attempt and completion.** The table presents the results of three separate multivariable logistic regression models, using the same set of independent variables to predict consent, battery attempt, and completion. Wald test P-values reflect the statistical significance of each variable. Odds ratios (OR), P-values (P), and 95% confidence intervals (95% CI) were provided for each category.

|  | **Consent** | | | | **Attempt** | | | | **Complete** | | | |
| --- | --- | --- | --- | --- | --- | --- | --- | --- | --- | --- | --- | --- |
| **Variable** | **OR** | **P** | **95% CI** | **Wald test P** | **OR** | **P** | **95% CI** | **Wald test P** | **OR** | **P** | **95% CI** | **Wald test P** |
| **Sex** |  |  |  |  |  |  |  |  |  |  |  |  |
| Female | Reference | | | | | | | | | | | |
| Male | 0.79 | 0.11 | 0.59, 1.06 |  | 1.04 | 0.84 | 0.68, 1.62 |  | 0.51 | **0.03** | 0.28, 0.94 |  |
| **Handedness** |  |  |  |  |  |  |  |  |  |  |  |  |
| Right | Reference | | | | | | | | | | | |
| Left | 0.68 | 0.11 | 0.43, 1.08 |  | 0.94 | 0.88 | 0.44, 2.01 |  | 0.39 | **0.04** | 0.71, 0.93 |  |
| **Education** |  |  |  | **0.046** |  |  |  | 0.06 |  |  |  | 0.16 |
| Non-attempted | Reference | | | | | | | | | | | |
| Below O-levels | 0.89 | 0.69 | 0.51, 1.55 |  | 4.22 | 0.01 | 1.35, 13.2 |  | 1.75 | 0.39 | 0.49, 6.23 |  |
| O-level | 1.40 | 0.10 | 0.93, 2.10 |  | 1.51 | 0.18 | 0.84, 2.75 |  | 1.50 | 0.37 | 0.61, 3.63 |  |
| A-level | 1.43 | **0.07** | 0.97, 2.12 |  | 1.84 | 0.04 | 1.02, 3.29 |  | 2.58 | 0.04 | 1.05, 3.63 |  |
| Degree | 2.10 | **0.01** | 1.23, 3.60 |  | 2.12 | 0.05 | 0.99, 4.53 |  | 3.55 | 0.03 | 1.17, 10.70 |  |
| **Childhood** **SEP** | |  |  |  |  |  |  |  |  |  |  |  |
| Manual | Reference | | | | | | | | | | | |
| Non-manual | 0.86 | 0.31 | 0.65, 1.15 |  | 1.49 | 0.06 | 0.98, 2.27 |  | 0.75 | 0.36 | 0.40, 1.39 |  |
| **Adult SEP** |  |  |  |  |  |  |  |  |  |  |  |  |
| Manual | Reference | | | | | | | | | | | |
| Non-manual | 1.38 | **0.07** | 0.43, 1.09 |  | 1.01 | 0.98 | 0.57, 1.79 |  | 1.08 | 0.86 | 0.47, 2.46 |  |
| **Childhood cognitive score** | | | | 0.10 |  |  |  | 0.26 |  |  |  | 1.00 |
| Bottom 10% | Reference | | | | | | | | | | | |
| Middle 80% | 1.60 | 0.048 | 1.00, 2.59 |  | 1.81 | 0.11 | 0.88, 3.97 |  | 0.00 | nan | nan, nan |  |
| Top 10% | 2.96 | 0.046 | 1.01, 3.82 |  | 1.63 | 0.32 | 0.63, 4,26 |  | 0.00 | nan | nan, nan |  |
| **Word learning test memory score at age 69** | | | | 0.003 |  |  |  | 0.52 |  |  |  | 0.68 |
| Bottom 10% | Reference | | | | | | | | | | | |
| Middle 80% | 2.86 | 0.003 | 1.27, 3.10 |  | 0.59 | 0.28 | 0.23,1.54 |  | 1.56 | 0.10 | 0.29, 3.42 |  |
| Top 10% | 1.98 | 0.001 | 1.52, 5.42 |  | 0.68 | 0.51 | 0.22, 2.12 |  | 1.00 | 0.58 | 0.33, 7.32 |  |
| **Mental health prevalence at age 69** | | | |  |  |  |  |  |  |  |  |  |
| No | Reference | | | | | | | | | | | |
| Yes | 0.89 | 0.57 | 0.58, 1.35 |  | 1.17 | 0.65 | 0.59, 2.32 |  | 0.92 | 0.87 | 0.34, 2.46 |  |
| **Lifetime smoking by 69 years** | | | | 0.08 |  |  |  | 0.26 |  |  |  | **0.048** |
| Never smoke | Reference | | | | | | | | | | | |
| Ex-smoker | 1.18 | 0.26 | 0.89, 1.57 |  | 1.16 | 0.24 | 0.75, 1.80 |  | 1.04 | 0.91 | 0.54, 1.97 |  |
| Current smoker | 0.63 | 0.14 | 0.34, 1.16 |  | 0.57 | 0.50 | 0.22, 1.45 |  | 0.19 | 0.02 | 0.05, 0.78 |  |
| **Alcohol use at age 69** | | | | **0.04** |  |  |  | **0.02** |  |  |  | 0.72 |
| Never | Reference | | | | | | | | | | | |
| Less than once a week | 1.40 | 0.23 | 1.16, 3.49 |  | 1.73 | 0.2 | 0.75, 4.01 |  | 1.53 | 0.53 | 0.39, 5.87 |  |
| 2-3 x per week | 1.75 | **0.05** | 1.01, 3.03 |  | 3.42 | **0.01** | 1.40, 8.33 |  | 1.49 | 0.56 | 0.39, 5.64 |  |
| 4+ per week | 2.02 | **0.01** | 1.16, 3.49 |  | 1.70 | 0.21 | 0.73, 3.94 |  | 1.96 | 0.32 | 0.52, 7.39 |  |
| **Weight status at age 69** | | | | 0.45 |  |  |  | **0.04** |  |  |  | 0.99 |
| Normal | Reference | | | | | | | | | | | |
| Overweight | 0.82 | 0.21 | 0.59, 1.12 |  | 1.01 | 0.98 | 0.61, 1.63 |  | 0.99 | 0.97 | 0.50, 1.93 |  |
| Obese | 0.87 | 0.46 | 0.61 ,1.25 |  | 0.57 | **0.04** | 0.33, 0.97 |  | 0.99 | 0.97 | 0.44, 2.23 |  |
| **Overall disease burden at age 69** | | | | 0.59 |  |  |  | 0.58 |  |  |  | **0.04** |
| 0 | Reference | | | | | | | | | | | |
| 1 | 1.24 | 0.22 | 0.89, 1.73 |  | 0.79 | 0.37 | 0.47, 1.32 |  | 0.30 | 0.01 | 0.13, 0.72 |  |
| 2 | 1.03 | 0.88 | 0.70, 1.52 |  | 1.00 | 1.00 | 0.53, 1.88 |  | 0.37 | 0.04 | 0.14, 0.96 |  |
| 3 and 3+ | 1.67 | 0.77 | 0.68, 1.67 |  | 0.69 | 0.27 | 0.35, 1.34 |  | 0.25 | 0.01 | 0.09, 0.75 |  |
| **Self-rated health at age 68** | | | | 0.01 |  |  |  | 0.95 |  |  |  | 1.00 |
| Poor | Reference | | | | | | | | | | | |
| Fair | 1.47 | 0.61 | 0.33, 6.69 |  | 1.27 | 0.85 | 0.10, 16.3 |  | 0.00 | nan | nan, nan |  |
| Good | 1.97 | 0.37 | 0.44, 8.67 |  | 1.47 | 0.76 | 0.12, 17.6 |  | 0.00 | nan | nan, nan |  |
| Very good | 2.51 | 0.23 | 0.57, 11.13 |  | 1.47 | 0.76 | 0.12, 17.8 |  | 0.00 | nan | nan, nan |  |
| Excellent | 4.17 | 0.07 | 0.90, 19.49 |  | 1.82 | 0.64 | 0.14, 23.1 |  | 0.00 | nan | nan, nan |  |

SEP=socioeconomic position

Consent: Number of observations=1137, Pseudo R-squared=0.09, LLR p < 0.001

Attempt: Number of observations= 724, Pseudo R-squared=0.08, LLR p < 0.05

Completion: Number of observations=585, Pseudo R-squared=0.10, LLR p=0.04

**Supplementary Table 5: Separate univariate regression for the completion of online battery.**

| **Variable** | **OR** | **P** | **95% CI** |
| --- | --- | --- | --- |
| **Sex** |  |  |  |
| Female | Reference | | |
| Male | 0.73 | 0.15 | 0.47, 1.12 |
| **Handedness** |  |  |  |
| Right | Reference | | |
| Left | 0.39 | 0.004 | 0.21, 0.74 |
| **Education** |  |  |  |
| Non-attempted | Reference | | |
| Below O-levels | 2.30 | 0.15 | 0.74, 7.17 |
| O-level | 1.6 | 0.15 | 0.84, 3.04 |
| A-level | 2.17 | 0.02 | 1.16, 4.07 |
| Degree | 2.37 | 0.02 | 1.12, 5.01 |
| **Childhood** **SEP** |  |  |  |
| Manual | Reference | | |
| Non-manual | 1.04 | 0.88 | 0.65, 1.64 |
| **Adult SEP** |  |  |  |
| Manual | Reference | | |
| Non-manual | 1.07 | 0.84 | 0.57, 1.99 |
| **Childhood cognitive score** | |  |  |
| Bottom 10% | Reference | | |
| Middle 80% | 0.20 | 0.11 | 0.03, 1.44 |
| Top 10% | 0.18 | 0.11 | 0.02, 1.43 |
| **Word learning test memory score at age 69** | | |  |
| Bottom 10% | Reference | | |
| Middle 80% | 1.26 | 0.64 | 0.48, 3.35 |
| Top 10% | 2.33 | 0.19 | 0.67 ,8.13 |
| **Mental health prevalence at age 69** | | |  |
| No | Reference | | |
| Yes | 0.96 | 0.93 | 0.44, 2.09 |
| **Lifetime smoking by 69 years** | | |  |
| Never smoke | Reference | | |
| Ex-smoker | 0.87 | 0.60 | 0.52, 1.45 |
| Current smoker | 0.28 | 0.01 | 0.11, 0.74 |
| **Alcohol use at age 69** | | |  |
| Never | Reference | | |
| Less than once a week | 1.27 | 0.68 | 0.40, 4.00 |
| 2-3 x per week | 1.27 | 0.68 | 0.41, 3.95 |
| 4+ per week | 1.34 | 0.61 | 0.43, 4.14 |
| **Weight status at age 69** | | |  |
| Normal | Reference | | |
| Overweight | 0.72 | 0.26 | 0.40,1.28 |
| Obese | 0.73 | 0.36 | 0.38,1.42 |
| **Overall disease burden at age 69** | | |  |
| 0 | Reference | | |
| 1 | 0.27 | 0.06 | 0.27, 1.04 |
| 2 | 0.25 | 0.13 | 0.25, 1.19 |
| 3 and 3+ | 0.21 | 0.06 | 0.21, 1.04 |
| **Self-rated health at age 68** | | |  |
| Poor | Reference | | |
| Fair | 1.70 | 1.00 | 0.00, inf |
| Good | 2.42 | 1.00 | 0.00, inf |
| Very good | 2.43 | 1.00 | 0.00, inf |
| Excellent | 3.56 | 1.00 | 0.00, inf |

SEP=socioeconomic position

**Supplementary Table 6: Logistic regression models on first task performance for battery completion**

Number of observations = 813, Pseudo R-squared = 0.008, LLR p-value = 0.03

| **Variable** | **P** | **OR** | **95% CI** | |
| --- | --- | --- | --- | --- |
| Summary score | 0.03 | 1.03 | 1.00 | 1.05 |

**Supplementary Table 7: Description of emerging themes from the Qualitative Analysis**

| **Theme** | | **Description** | |
| --- | --- | --- | --- |
| **Technical Issues** | | Broad theme for all technical issues related to the website, links or anything else. | |
| **Subcategories** | | **Description** | **Significant statement examples** |
| Issues accessing link | | Any technical issues related to the links to either Cognitron platform, consent form or information sheet. | |
|  | Copy and paste | Participant refers to issues copying and pasting. | *Very sorry but I am not technically versed so having filled in the appropriate details I am now not able to continue as I can’t copy and paste as requested as I don’t know how.* |
|  | Requests resend link | Participant is unable to access the link and request it is resent. | *I have completed the Consent Form but somehow have lost the link to the actual Questionnaire__ please can you resend to me?* |
|  | Return to link | Any issues returning to Cognitron platform. | *When I returned there was a message saying that my time had expired and I could not get back in to retrieve the link.* |
| Technical issues with Cognitron platform | | Any technical issues that refer to the Cognitron platform, exclusive of accessing the link. | |
|  | Not responding | Participant notes that the Cognitron platform is not working as it should. | *I attempted to complete the thinking tests but unfortunately it doesn’t respond to my answers. Right from the start it just freezes. I know the internet connection is not great in my area, so that could be the problem.* |
|  | Rotation Lock | Reported issues with tablets not rotating to the correct orientation. | *Sorry I could not complete this. I could not get my iPad to rotate to portrait view.* |
| **General Queries** | | Any queries or questions from participants | |
| **Subcategories** | | **Description** | **Significant statement examples** |
| Follow-up queries | | A reason for contacting the helpful after completing the tests | |
|  | Request for results | Participant requests their results or requests to see how well they performed. | *Do we get the results of the tasks?* |
|  | Closing response | Participant replied to our response either confirming the issue was resolved or by providing more details about the problem. | *I have now completed the tasks. Sorry about the confusion.* |
| How to perform task | | Participant queries how to perform the task or ask for clarification on the instructions. | |
|  | Resubmission | Participant requests or informs us that they did not complete the tasks in one sitting. | *I just needed a break from the first set of tests. Am I expected to complete all four sections in one go?* |
|  | Device query | Participant asks what devices can be used. | *Can I do these with a Chromebook?* |
|  | Different form | Participant requests to complete or receive the tasks in a different form. | *Sorry but I’m not into computers and links but if you wish to send me a paper questionnaire.* |
| Privacy and security concerns | | Privacy and security concerns regarding the tasks or the links in the invitation email. | *The email below comes from an address that is not recognized. Is this a phishing scam?* |
| **Withdrawal Reason** | | Reason provided as to why participant withdrew from Online46. | |
| **Subcategories** | | **Description** | **Significant statement examples** |
| Not available | | Participant is unable to or unavailable to participate during the timeframe of the study. | |
|  | Too busy | Unable to complete the tasks as they are too busy. | *I am very busy and cannot spare the time at present as I have other priorities.* |
|  | Personal or family matter | Unable to complete the tasks due to a personal or family matter. | *I am now in a position of maybe loosing my accommodation and am very sorry but can’t concentrate on anything else until this is sorted.* |
|  | On holiday | Unable to complete the tasks due to being on holiday and not returning before the deadline. | *Sorry, but at the moment I am out of the country on holiday.* |
| Technology access | | Participant is unable to participate due to access to appropriate technology. | *I was unable to access the patient information link or consent form, due to the age of this laptop.* |
| Personal health or death | | Participant is unable to participate due to personal health or death. | *I regret that I can’t complete the task. I am recovering from an operation and not able to do it.* |
| None given | | No reason given for not wishing to participate in study. | *Thank you, but would prefer not to participate at the moment.* |
| Dislike cognitive tasks | | Participant refers to not liking memory and thinking tasks. | *The wretched experience of the last task requested has put me off.* |
| **Feedback** | | Any feedback from participants | |
| **Subcategories** | | **Description** | **Significant statement examples** |
| Positive feedback | | Participant comments said they either enjoyed the tasks or the study. | *Some of them were great fun.* |
| Suggestions | | Participants offer their feedback on either how the instructions are worded, the tasks themselves or the platform. | *I was not able to read the instructions (too small and faint) or see the symbols easily. I could not enlarge them.* |
| Did not understand taks | | Participants referenced that they did not understand the instructions or did not know how to respond. | *I started your task and on reaching blocks I struggled to understand what was required.* |
| Negative feedback | | Participant makes a comment that they did not enjoy the tasks or that these tasks caused a negative reaction. | *Cannot continue as it causes an unpleasant reaction. Hot flushes and sweating caused by the mild stress.* |
| **Subjective reports** | | Commentary from participants on their performance or how they performed the task. | |
| **Subcategories** | | **Description** | **Significant statement examples** |
| Lack of tech literacy | | Participant makes a remark that they lack technical literacy. | *I’m not anti-questionnaires if they appear on paper, but modern on-line technology is beyond me. I wonder what percentage of the geriatrics on your survey have the technology or the skill to use it to manage these electronic surveys.* |
| Cognitive performance | | Participant makes a comment on how they performed on the test or Participant provided a reason they have performed sub-optimally. | *I actually find the memory tasks the most stressful of all the MRC project’s tasks. I have always had a very good memory – at least a very good long-term memory, but, of course I know my memory, and particularly my short term memory, is deteriorating quiet fast, and I don’t like to be made really conscious of this by doing the tasks.* |

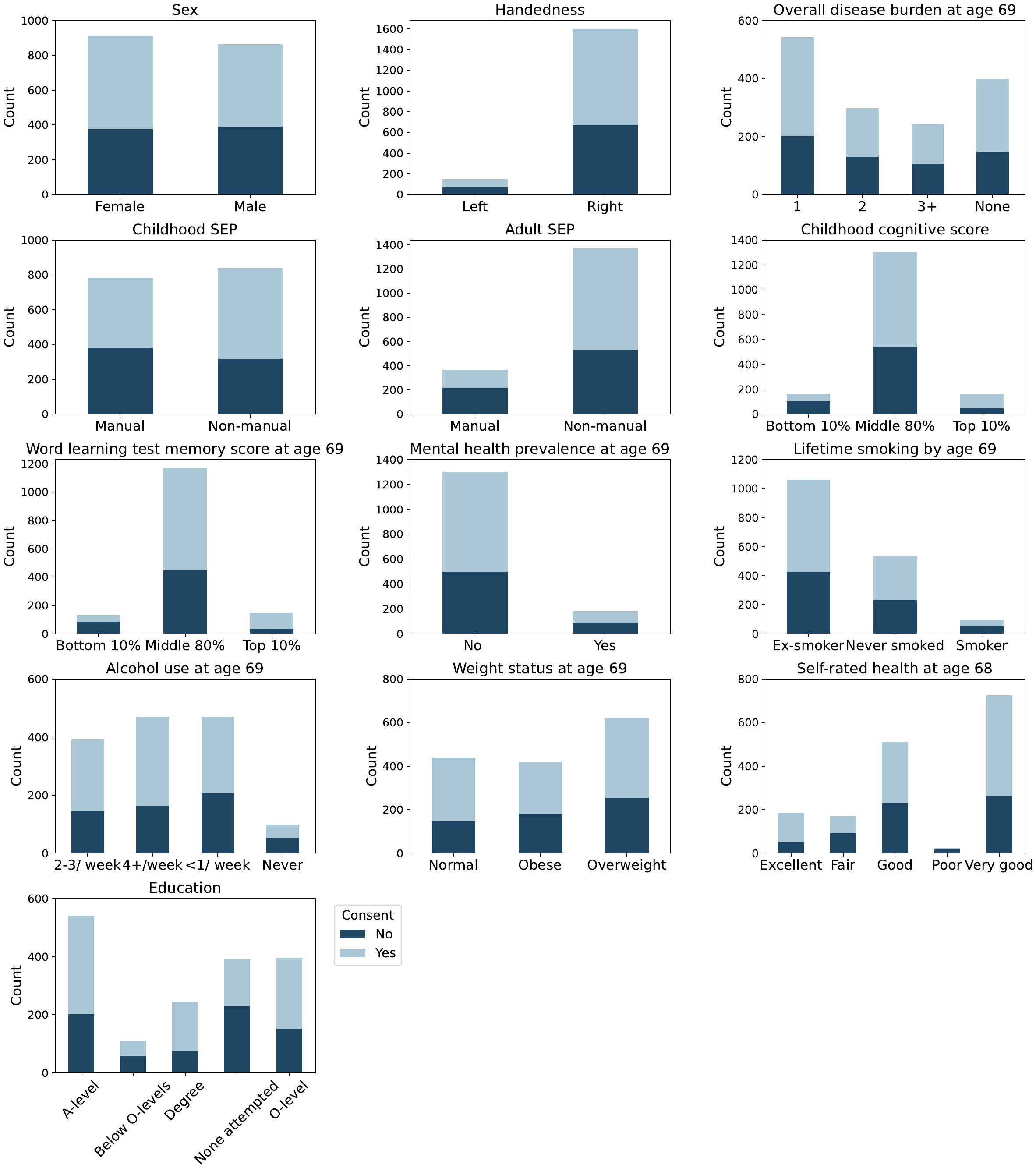

**Supplementary Figure 1 - Distribution of sociodemographic factors and health features for the consent of the online battery.**  The stacked bar plots show the number of NSHD members who consented or did not consent to conduct the online cognitive tasks for each category of a specific variable.

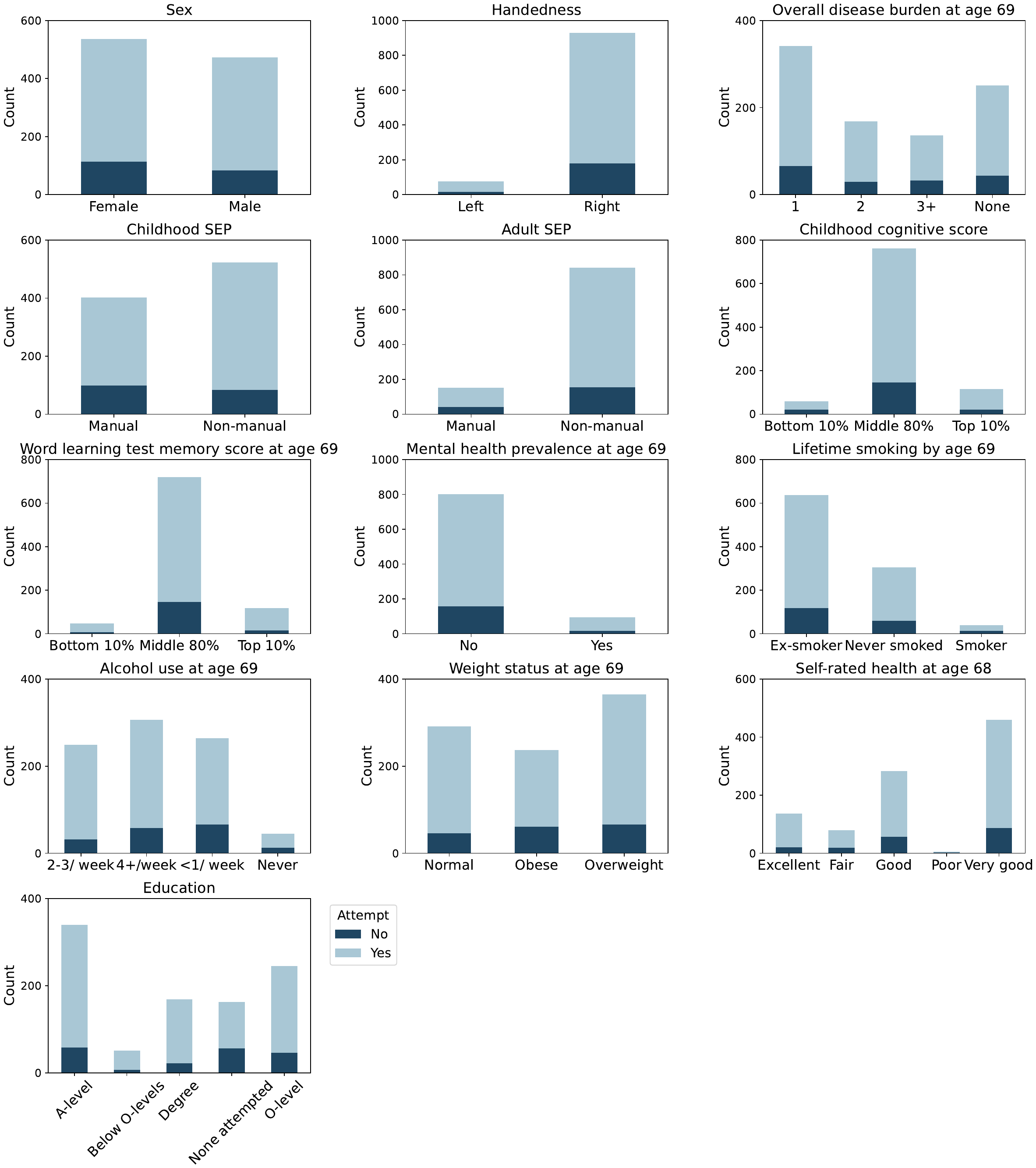

**Supplementary Figure 2 - Distribution of sociodemographic factors and health features for the attempt of online battery.** The stacked bar plots show the number of consented members who attempted or did not attempt the online cognitive tasks for each category of a specific variable.

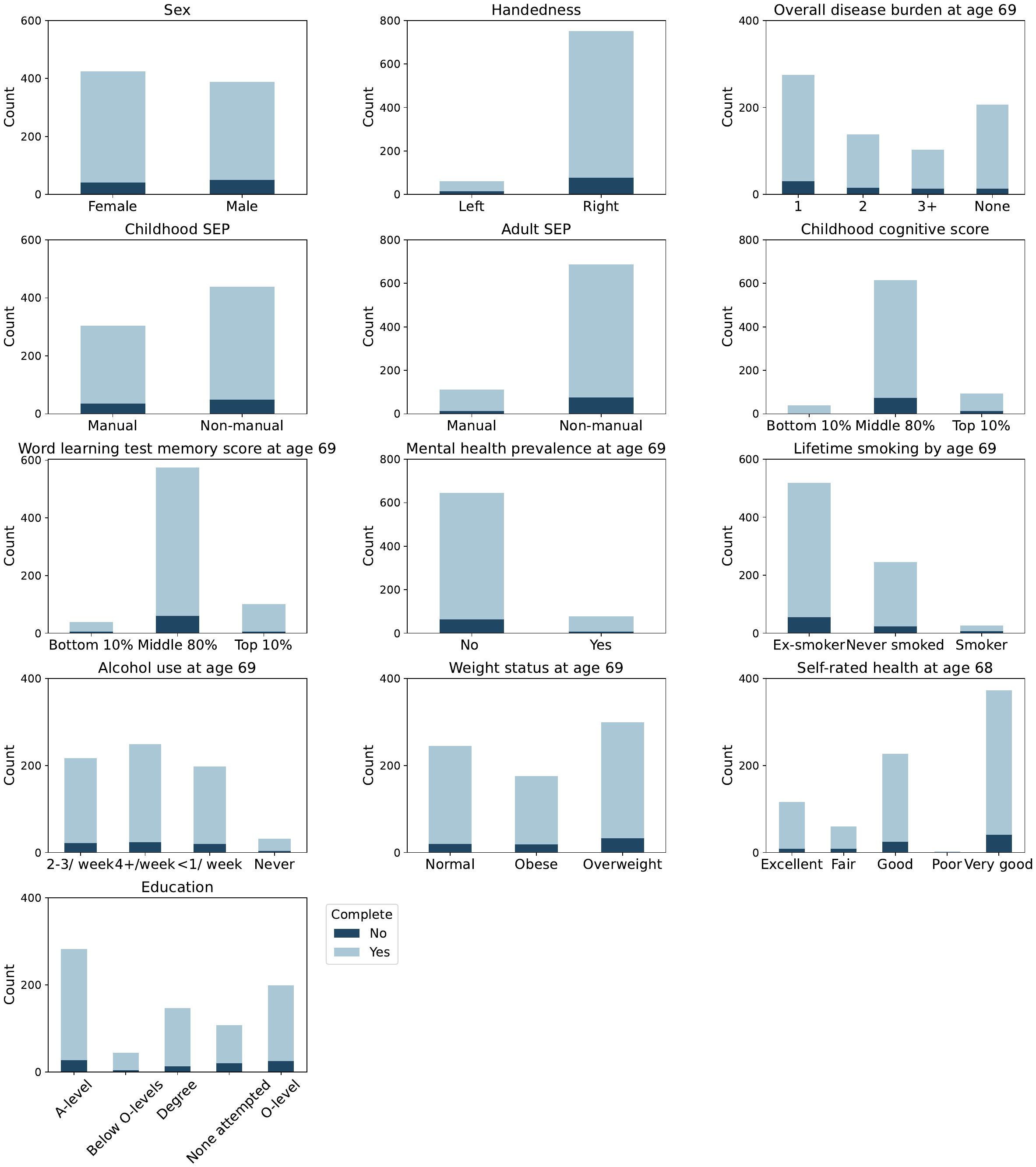

**Supplementary Figure 3 - Distribution of sociodemographic factors and health features for completion of the online battery.** The stacked bar plots show the number of members who attempted the battery and either completed or did not complete the online cognitive tasks for each category of a specific variable.

**Supplementary Text 1: Description of the automatic method to derive the lower summary score threshold for each task**

Summary scores different from 0 (indicating potentially skipped or unanswered questions) are extracted. The filtered scores are then sorted in ascending order, and the 95% quantile of the score distribution is identified. This 95% quantile is divided by a user-defined parameter to determine the number of bins for segmenting the score distribution, with a larger divisor resulting in less stringent filtering. Starting from the leftmost bin, the function compares bin heights. If a subsequent bin height is lower than the previous one, values in the preceding bin are flagged as unreliable. This process continues until reaching the 5% quantile of the summary score distribution. The maximum value of the last bin flagged as unreliable is set as the summary score threshold. Values below this threshold are considered too low to be reliable.
